# Real world validation of activity recognition algorithm and development of novel behavioral biomarkers of falls in aged control and movement disorder patients

**DOI:** 10.1101/2022.11.23.22282685

**Authors:** Ali Nouriani, Alec Jonason, Luke Sabal, Jacob Hanson, James Jean, Thomas Lisko, Emma Reid, Yeng Moua, Shane Rozeboom, Kaiser Neverman, Casey Stowe, Rajesh Rajamani, Robert A. McGovern

**Affiliations:** Laboratory for Innovations in Sensing, Estimation and Control, Department of Mechanical Engineering, College of Science and Engineering, University of Minnesota, Minneapolis, MN, USA; Department of Neurosurgery, University of Minnesota Medical School, Minneapolis, MN, USA; Rocky Vista University College of Osteopathic Medicine, Parker, CO, USA; Division of Neurosurgery, Minneapolis Veterans Affairs Health Care System, Minneapolis, MN, USA

**Keywords:** Falls, Near-falls, Postural Instability, Gait, Parkinson disease, Wearable sensors

## Abstract

The use of wearable sensors in movement disorder patients such as Parkinson’s disease (PD) and normal pressure hydrocephalus (NPH) is becoming more widespread, but most studies are limited to characterizing general aspects of mobility using smartphones. There is a need to accurately identify specific activities at home in order to properly evaluate gait and balance at home, where most falls occur.

We developed an activity recognition algorithm to classify multiple daily living activities including high fall risk activities such as sit to stand transfers, turns and near-falls using data from 5 inertial sensors placed on the chest, upper-legs and lower-legs of the subjects. The algorithm is then verified with ground truth by collecting video footage of our patients wearing the sensors at home.

Our activity recognition algorithm showed >95% sensitivity in detection of activities. Extracted features from our home monitoring system showed significantly better correlation (∼69%) with prospectively measured fall frequency of our subjects compared to the standard clinical tests (∼30%) or other quantitative gait metrics used in past studies when attempting to predict future falls over one year of prospective follow-up.

Although detecting near-falls at home is difficult, our proposed model suggests that near-fall frequency is the most predictive criterion in fall detection through correlation analysis and fitting regression models.

## 1 Introduction

Postural instability is both a cardinal symptom of movement disorders like Parkinson’s disease (PD) and a major cause of falls in these patients [1]. Injurious falls and hip fractures occur at higher rates in PD patients 10-15 years prior to diagnosis of PD compared to healthy controls [1]. Therefore, if subtle balance dysfunction could be properly identified and characterized, this information could be used to initiate falls preventions and physical therapy programs, improve fall prediction algorithms, and monitor or evaluate new treatments.

The Movement Disorder Society-Unified Parkinson’s Disease rating scale (MDS-UPDRS) [2], [3] analyzes all motor symptoms using a semi-quantitative scale. Its validity and reliability is well recognized and it is the clinical gold-standard in terms of monitoring symptoms related to PD [4]. However, these assessments are subject to inter-rater variability, and the unavailability of continuous monitoring limits these methods. The score of the evaluation depends on the patient’s current status, which may fluctuate day-to-day and depending on the time since the last dose of medication was taken. On the other hand, traditional lab-based assessments using infrared cameras or quantitative analysis to characterize postural instability in patients with movement disorders are costly, not portable, and are unable to track long-term movement data from these patients in their day-to-day lives when most falls occur. Therefore, there is a serious need for long-term, real-time, and objective characterization of movement as a complement to clinical and lab-based assessments [7].

A few methods of characterizing mobility in patients with movement disorders have been proposed, including home movement diaries [5] and characterizing mobility using smartphone applications. Patient diaries and questionnaires at home are frequently used in clinical routine to study motor stages and fluctuations in late-stage PD [6]. However, diaries are subject to fatigue, errors, and bias which impacts the quality and credibility of the data, particularly in patients with cognitive dysfunction [6]. Some methods of tracking participants at home may involve using mobile phone-based systems that gather data using inertial sensors that are built into smartphones [8]. This yields data that allows for general tracking of activities such as walking, sitting and sleeping, but does not provide quantitative insights into participants’ postural responses when experiencing a fall or near-fall. Additionally, relying on data from a device that is not fixed to the patient’s body may introduce error or leave long gaps in data. More elaborate systems using multiple cameras throughout a person’s home in order to track their movements may also be used, but this may not be feasible on a wide scale due to its cost, complexity, and privacy concerns [9].

To address some of these problems, we have developed an algorithm that can enable real-time detection of high-risk activities in the patient’s home environment using inexpensive and widely available wearable technology. The goal of this study was to create a video-validated dataset of movement disorder patients and healthy controls engaged in daily living activities in their homes, develop an algorithm for automatic recognition of near-falls/high fall risk activities and subsequently quantitatively characterize the patient’s response to these events. Finally, we developed novel behavioral biomarkers based on this data to assess their relationship to patients’ prospective fall risk over one year of follow-up.

## 2 Methods

### 2.1 Participant population characteristics

19 movement disorder patients who were being clinically evaluated and/or treated for either normal pressure hydrocephalus or Parkinson’s disease and 10 age-matched healthy control participants were enrolled over a period of 2 years from the Minneapolis VA Health Care System (MVAHCS) and University of Minnesota (UMN). Enrollment was designed to enroll a variety of types of movement disorder patients with varying gait dysfunction and postural instability ranging from normal gait and balance (MDS-UPDRS gait and pull test item scores of 0) to moderate dysfunction (MDS-UPDRS scores of 3). Patient participants were excluded if they were non-ambulatory or if they were unable to give consent. Control participants were excluded if they had any movement, gait, or balance disorders. Demographic information was collected (Table 1). This study was approved by the MVAHCS and UMN Institutional Review Boards, and all participants provided informed consent for participation according to the Declaration of Helsinki.

**Table 1.** Demography of participants

| Condition<br>* | Age<br>[year] | Female<br>*n*, % | Height<br>[cm] | Weight<br>[kg] | UPDRS<br>Pull test | UPDRS<br>Gait | Duration<br>of Disease<br>[year] |
| --- | --- | --- | --- | --- | --- | --- | --- |
|  | *Mean*<br>(*SD*) |  | *Mean*<br>(*SD*) | *Mean*<br>(*SD*) | *Mean*<br>(*SD*) | *Mean*<br>(*SD*) | *Mean*<br>(*SD*) |
| **PD**<br>(**n = 11**) | 65 (4.76) | 2, 20% | 181.54<br>(8.97) | 92.94<br>(23.63) | 0.56 (0.96) | 0.90 (0.74) | 10.50<br>(5.41) |
| **NPH**<br>(**N = 8**) | 69.8 (8.21) | 1, 11% | 180.72<br>(3.50) | 102.57<br>(20.71) | 1.04 (0.56) | 1.85 (0.89) | N/A |
| **C**<br>(**n = 10**) | 61.1 (9.97) | 8, 80% | 172.20<br>(6.83) | 79.75<br>(27.69) | 0<br>(0) | 0<br>(0) | N/A |

\* PD = Parkinson's Disease, NPH = Normal pressure hydrocephalus, C = Control

### 2.2 Measurement setup

The measurement sensors were customized and reprogrammed inertial measurement units (IMU; SparkFun, Inc. Boulder, CO, USA). The board was equipped with a high-performance ARM Cortex-M4 processor powered by 500 mAh high-capacity Lithium battery [10]. The measurement Integrated circuit (IC) was an ICM-20948 (InvenSense, San Jose, CA, USA) which can log nine degrees of freedom (accelerometer, gyroscope, magnetometer) at nearly 250 Hz [11]. The data from the IMU was sampled with a 100Hz frequency and stored on a flash memory though it can be streamed wirelessly through Bluetooth connectivity to a smartphone or computer (Figure 1).

**Fig 1.**
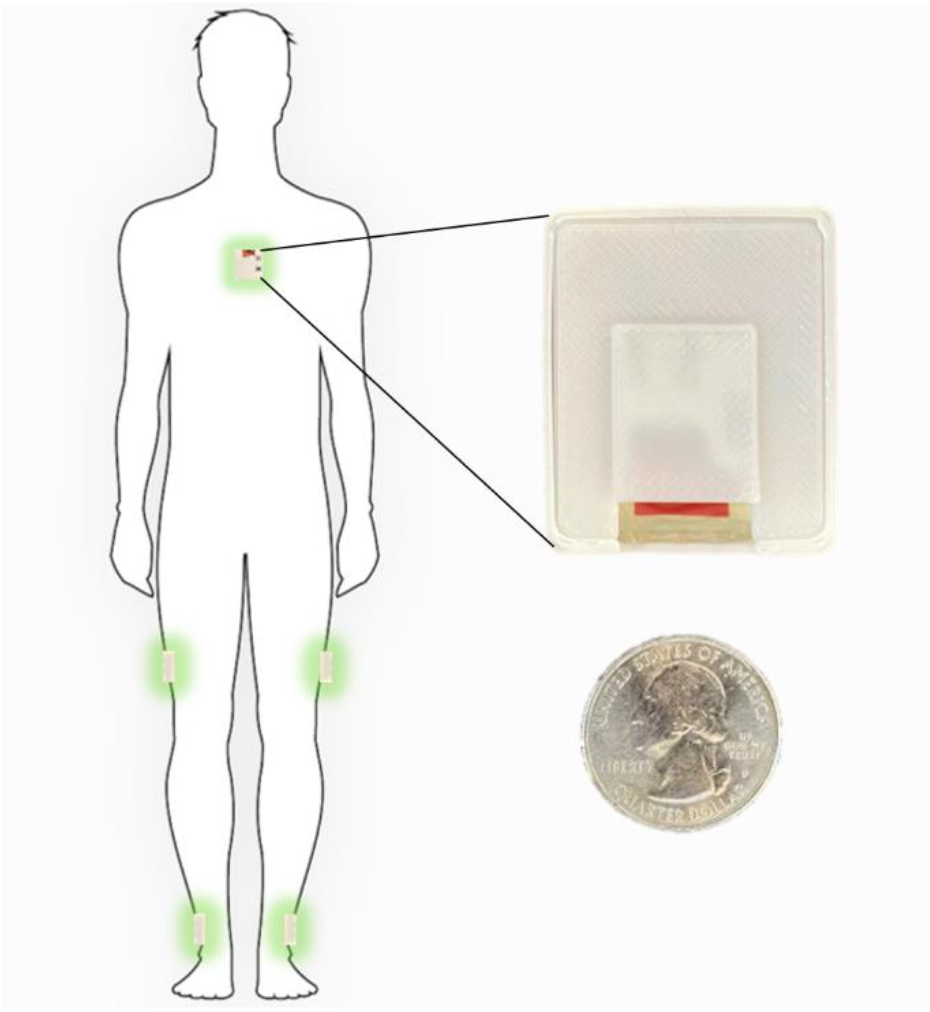
Inertial sensors placement on the body in 5-sensor configuration, one on the chest and one on each lower leg and one on each upper leg.

The sensor configuration is one IMU sensor on each shank (just above the ankle), one IMU on each thigh, and one IMU sensor on the chest. This five sensor configuration uses an angle-based method taking advantage of the geometry of human in-plane walking. The details of our estimation algorithm and the accuracy of a variety of kinematic variables which can be calculated using this configuration compared to a gold standard infrared camera measurements has been previously described [16].

### 2.3 Home Wearable Sensor Usage

Each participant was shown how to properly place the IMUs (Figure 1) in the clinic at their baseline visit. They were then sent home for one week and were instructed to wear the IMUs during all waking hours. The IMUs were charged overnight. The entirety of the dataset was able to be stored on the available flash memory on the sensor board and therefore participants did not need to upload data or stream any data to an app. They simply wore the sensors during the day and charged them at night. Each patient is provided with a custom charger connected to a Raspberry Pi Zero board (Raspberry Pi Foundation, Cambridge, England) which is programmed to synchronize the sensors together using threading with an extremely accurate real-time clock module (DS3231 RTC, Adafruit, New York, NY) every time the sensors are connected to the charger [14]. During the week of wearable sensor use, a research coordinator contacted the participants daily to troubleshoot any technical problems and check in. Participants were then prospectively followed for one year and asked to complete fall diaries according to accepted fall data formatting. To supplement the fall diaries, a research coordinator called the participants weekly for the follow-up year to inquire about any falls occurring during the past week.

### 2.4 Home Wearable Sensor Activity Definitions and Video Validation

In order to properly identify home activities, we first defined a variety of activities using the IMU data (Supplemental Table 1). The algorithm used to identify each home activity is a deep learning-based activity recognition architecture using a convolutional neural network long short-term memory network (CNN-LSTM) which we have previously detailed [15]. We also used three other commonly used classifiers (logistic regression, support vector machine, decision tree) to compare their performance to our CNN-LSTM. The details of these algorithms can be found in the Supplemental Methods. In order to validate the algorithm-defined activities, we asked a subset of participants (n=10) to wear a small video camera (Runcam, Aberdeen, Hong Kong Island, Hong Kong, [12] with a necklace to wear at home. Each patient was asked to randomly record for 45-60 minutes each day, ideally while ambulating or performing some type of algorithm-detectable activity. The videos were then manually annotated using a video-defined equivalent of each IMU-defined home activity (Supplemental Table 1) and synchronized with the sensors using the camera timestamps. Examples of the video footage captured by the patients are provided in supplemental materials.

### 2.5 Fall Prediction Modeling

We prospectively followed all patient participants for one year with fall diaries and weekly individual participant contact to document the presence of any falls and the total number of falls over the entire year. From this data, we calculated the fall frequency as #falls/week. Using fall frequency as our outcome, we then examined the correlation between multiple computed features and fall frequency. These features ranged from standard demographic characteristics such as age, height, and weight, to clinical measurements such as the MDS-UPDRS pull test score, and also included a number of quantitative features from home measurements (denoted with an “_h”) that have been used in prior studies such as the total ambulatory time each day and number of ambulatory bouts each day (Supplemental Table 2). We also computed several novel features based on our previously video-validated activities described above. These include the frequency of near falls, turns and bends among others (events defined in Supplemental Table 1, all features used for correlation analysis are seen in Supplemental Table 2). We then created correlation confusion matrices to examine correlation of the previously mentioned data features with fall frequency. Using the fall diary data above, we also examined the time to first fall within the first year.

From the 29 participants, we have collected fall diaries and have survival data for 17 subjects. We did not collect the fall diaries from the control subjects and hence excluded them for the fall prediction model. From the remaining subjects, 9 are censored since they are either new patients, or their home data is missing, or stopped sending their fall diaries to us before week 52.

## 3 Results

### 3.1 Population characteristics

Eight patients with NPH and 11 with PD were enrolled for a total of 19 patient participants. Ten healthy, age-matched control participants were also enrolled. Demographic characteristics are demonstrated in Table 1. There were no significant differences in age, sex, height or weight between controls and patient participants. As expected, patient participants had significantly worse gait and postural stability MDS-UPDRS scores.

### 3.2 Activity Recognition Algorithm Validation

Ten of our patients generated more than 40 hours of video footage which was manually annotated (see Supplemental Table 1 for definitions). Figure 2 demonstrates an example of one day of recorded data using our sensors compared to the video footage obtained from a patient at home. This patient generated approximately 90 minutes of video footage during which he was ambulatory for approximately 45 minutes punctuated in the middle by 45 minutes where he was sitting at rest. The algorithm-predicted activity (blue) overlies the actual video-annotated activity (orange) for the vast majority of the time, with an example of one misclassified activity (∼8:58 am, standing misclassified as walking, seen in the Figure 2 inset).

**Fig 2.**
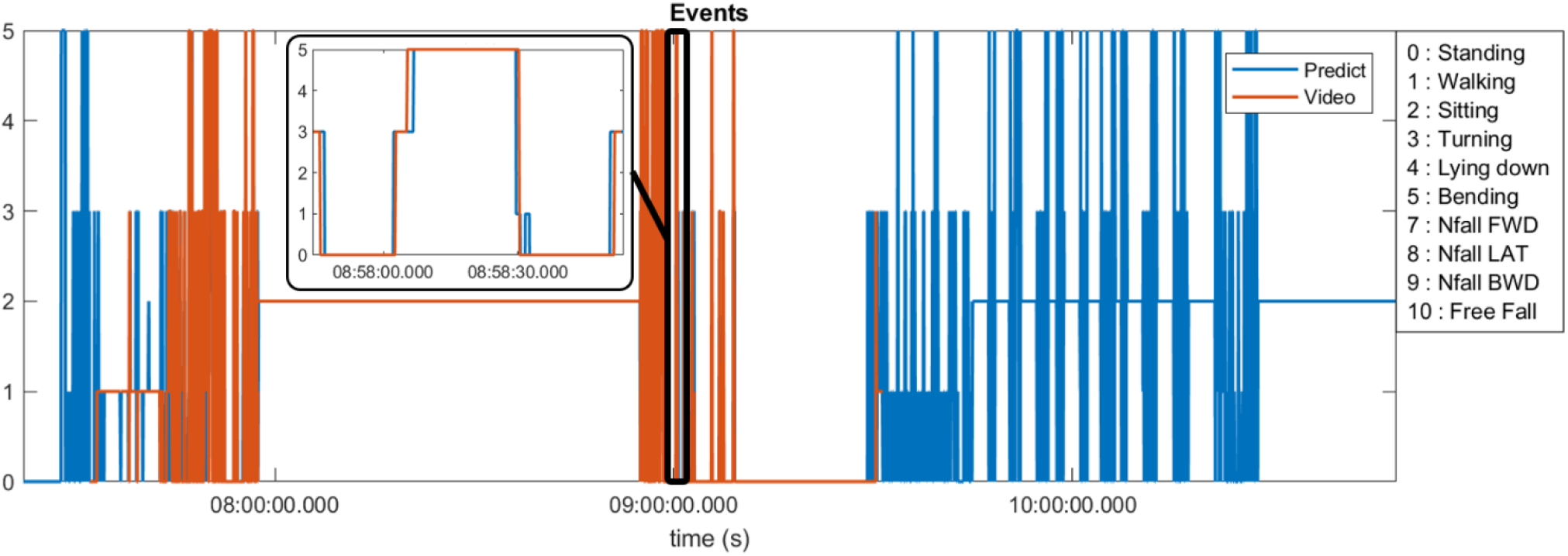
Predicted activities compared to the video annotations obtained from a patient at home environment

The sum total of video footage in the entire subset of patients resulted in more than 14,000 total events which were used for algorithm predictions. Figure 3 demonstrates the home activities predicted by our activity recognition algorithm in comparison to the video-annotated data collected on the subset of patients with video recorded events. Events which were common and straightforward to both define and detect such as walking, standing and turning demonstrated the highest accuracy (>99%). Because these three events were the most common overall, they also represented most of the false positive and false negative errors for all events. Bending, sitting and transitions from sit to stand or stand to sit were significantly less common and slightly less accurately predicted (91-94%). Near falls in any direction were among the least common events and were less accurately predicted (∼80%).

**Fig 3.**
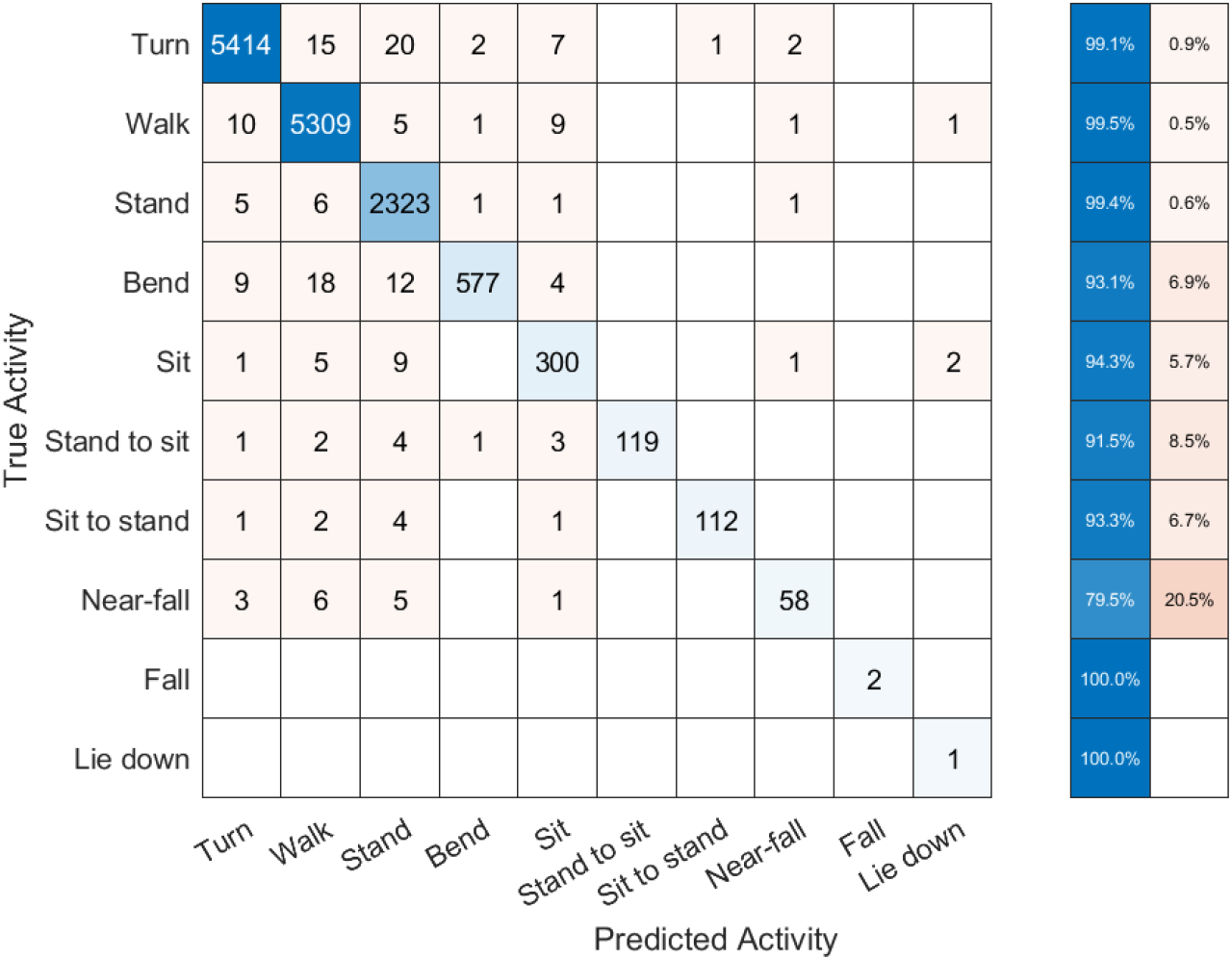
Confusion matrix of activity recognition algorithm results compared to the annotated videos for 6 subjects.

Table 2 shows the number of true positive (TP), false positive (FP), true negative (TN), and false negative (FN) samples from our activity recognition algorithm compared to the ground truth from video annotations. We can use these values to calculate the sensitivity (true positive rate or TPR), specificity (true negative rate or TNR), positive predictive value (PPV), negative predictive value (NPV), and accuracy (ACC). Because the number of total events is quite high, the specificity and overall accuracy of all the events are high, particularly for the low likelihood events such as sit to stand transitions, near-falls and falls. Nonetheless, even the low likelihood events had sensitivities >95% with the exception of near-falls which was 80%.

**Table 2.**
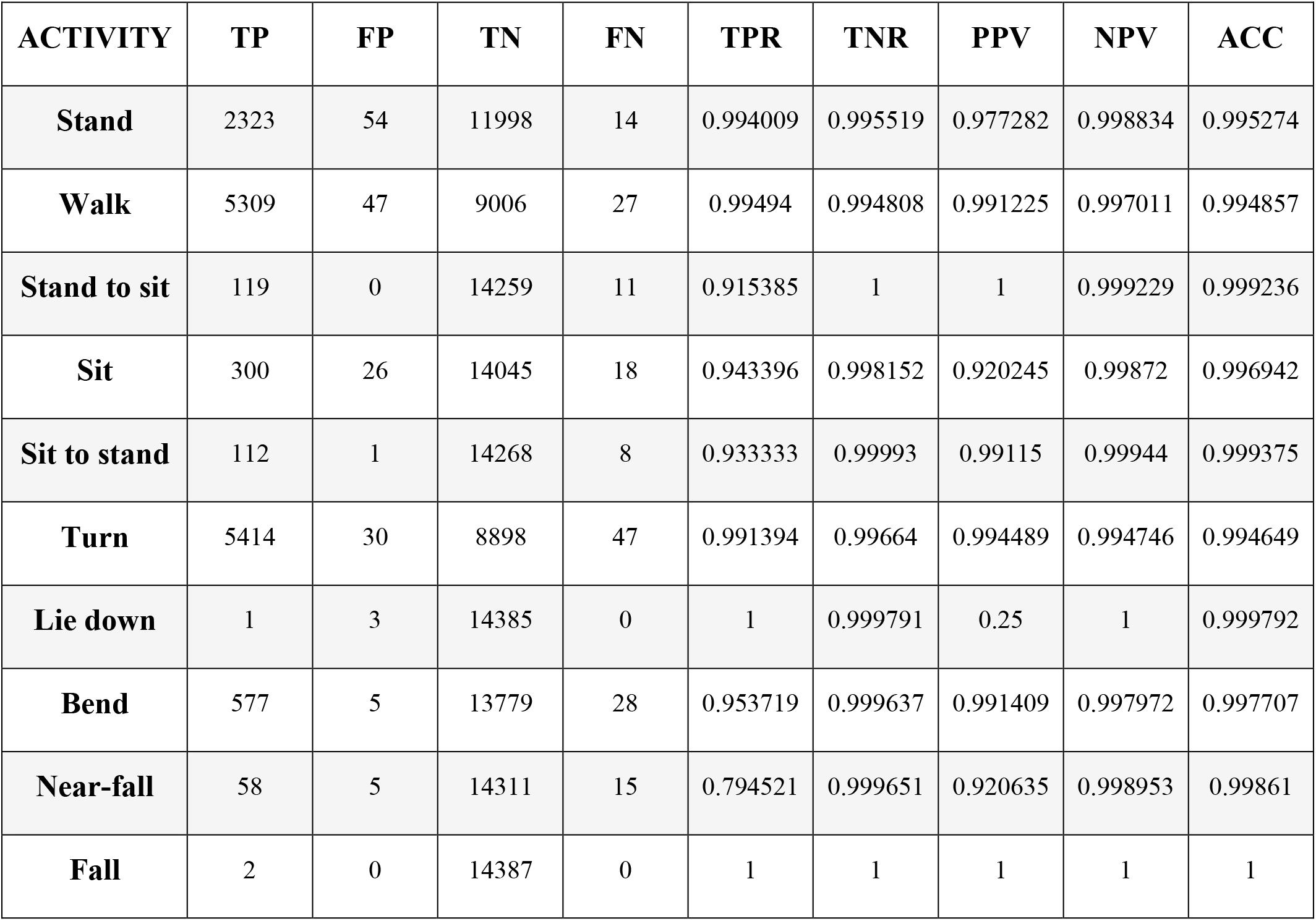
Statistics of each activity in our activity recognition algorithm

Supplemental figure 1 shows the median number of near-falls per week (Nfalls_h) for the three groups of our subjects. Control subjects show significantly fewernear-falls compared to PD and NPH patients. PD patients showed a slightly higher median compared to NPH patients which could be related to variations in the state of neurologicaldisease within these two groups. A detailed analysis and comparison across patient groups is out of scope of this paper and will be discussed in future publications.

To compare our LSTM algorithm to other standard classifiers commonly used to make predictions on large datasets, we have created receiver operating characteristic (ROC) curves for 6 activities of standing, walking, sit-stand transitions, turning, bending and near-falls. Figure 4 shows the ROC curves for four binary classifiers of Logistic Regression (LOG), Support Vector Machines (SVM), Decision Tree (DT), and our Long-Short-Term-Memory cells (LSTM) for each activity [15]. The area under the curve for each plot is summarized in Table 3. The performance of the LSTM classifier is superior in all activities with AUCs ranging from 0.982 to 0.999 for all activities while DT performs next best with slightly worse results. The SVM and LOG methods are significantly less accurate than LSTM or DT but still show acceptable results for a diagnostic classifier with AUCs ranging from 0.65 to 0.97. All classifiers worked well for relatively easily classified activities such as standing, walking or bending, but more difficult activities to classify such as near falls required the more sophisticated CNN-LSTM algorithm. There were no differences in activity classification between groups such that the CNN-LSTM algorithm was able to accurately classify activities in controls, NPH and PD patients equally well (Supplemental Figure 2). The overall performance (in terms of area under the curve) of each classifier is very similar across different patient groups. The small differences are related to random variables such as the number of samples, patient specific responses, and outlier responses.

**Table 3.** Area under the ROC curves for each activity

| CLASSIFIER | Standing | Walking | Sit-Stands | Turning | Bending | Near-Falls |
| --- | --- | --- | --- | --- | --- | --- |
| **LOG** | 0.85759 | 0.85745 | 0.65548 | 0.69863 | 0.96702 | 0.73180 |
| **SVM** | 0.99691 | 0.91850 | 0.71383 | 0.78733 | 0.95410 | 0.73401 |
| **DT** | 0.99999 | 0.99881 | 0.95043 | 0.95940 | 0.98589 | 0.90834 |
| **LSTM** | 0.99999 | 0.99985 | 0.98920 | 0.99288 | 0.99774 | 0.98253 |

**Fig 4.**
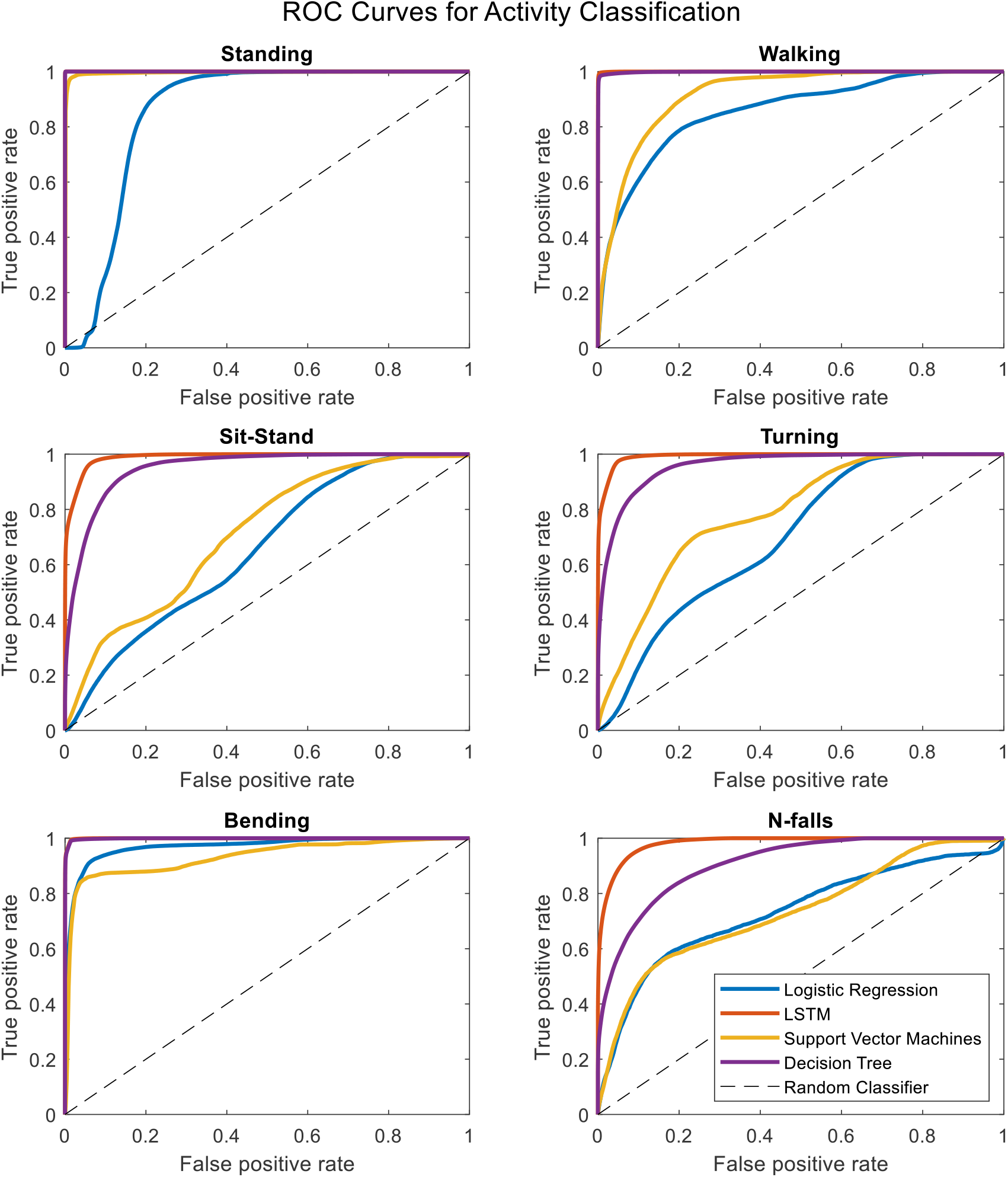
Receiver operating characteristic curves for four binary classifiers plotted separately for each activity.

Figure 5 demonstrates a correlation matrix examining the top ten features correlated with prospectively observed fall frequency over the subsequent year of follow-up. Two of our novel metrics, the total number and frequency of near falls detected by the home monitoring setup showed the highest correlation with patient fall frequency (0.69 and 0.67, respectively). Eight of the most correlated features with fall frequency come from values calculated solely using the IMUs while the patient moves around their home and surrounding environment. For example, time spent lying down per day (lie down frequency, i.e. lying duration/total time), the total number of ambulatory bouts at home (totNumABs), the frequency of sitting at home (sit_freq_h i.e sitting duration/total time), walking frequency at home (walk_freq_h), the peak acceleration of the chest at home (peak_acc_h, which represents the strongest perturbation the subject experiences at home as measured by the chest accelerometer in each day) and the distribution of ambulatory bouts in time (alpha parameter for ABs of more than 8 seconds (alpha_8)[17]) at home are all within the ten features with the highest correlation with fall frequency. It should be noted that some features were inversely correlated with fall frequency such that increased number of ambulatory bouts and increased walking frequency at home were associated with fewer falls (−0.40 and -0.36, respectively). Furthermore, Figure 5 also shows how correlated some of these features are with each other. For example, the total number of ABs are highly correlated with walking frequency (0.81), and the total number and frequency of near falls are almost perfectly correlated (0.98). We constructed a linear regression model on 5 features which are not highly correlated. The details of the regression model are presented in Tables 4 and 5. The model shows that near-fall frequency (nfall_freq), alpha parameter for ABs of more than 8 seconds (alpha_8) and UPDRS score (updrs) were the most significant predictors, respectively. The regression model parameters are summarized in Table 5. The only non-quantitative features included in the ten most correlated features were the MDS-UPDRS pull test item score measured in clinic, and the total number of failures (needing to be caught by examiner) in clinical pull tests (tot_failures). These were among the most weakly correlated features (0.30 and 0.26, respectively) overall and were not significantly associated with fall frequency in the multivariable linear regression model.

**Table 4.** Linear regression coefficients for fall frequency prediction

| Feature | Estimate | Standard error | t-stat | p-value |
| --- | --- | --- | --- | --- |
| **intercept** | 0.047251 | 0.020211 | 2.337934 | 0.041485 |
| **nfall_freq_h** | 0.696391 | 0.188534 | 3.69372 | 0.004151 |
| **totNumABs** | 6.82E-08 | 1.62E-06 | 0.042112 | 0.967239 |
| **sit_freq_h** | -0.00035 | 0.000178 | -1.99592 | 0.073889 |
| **updrs** | -0.01573 | 0.006449 | -2.43948 | 0.03488 |
| **alpha_8** | -0.00391 | 0.001185 | -3.30116 | 0.007998 |

**Table 5.** Linear regression model summary for fall frequency prediction

| Feature | SumSq | MeanSq | F | pValue |
| --- | --- | --- | --- | --- |
| **Total** | 0.014705 | 0.0009803 |  |  |
| **Model** | 0.012228 | 0.0024457 | 9.8737 | 0.001268 |
| **Residual** | 0.002477 | 0.0002477 |  |  |

**Fig 5.**
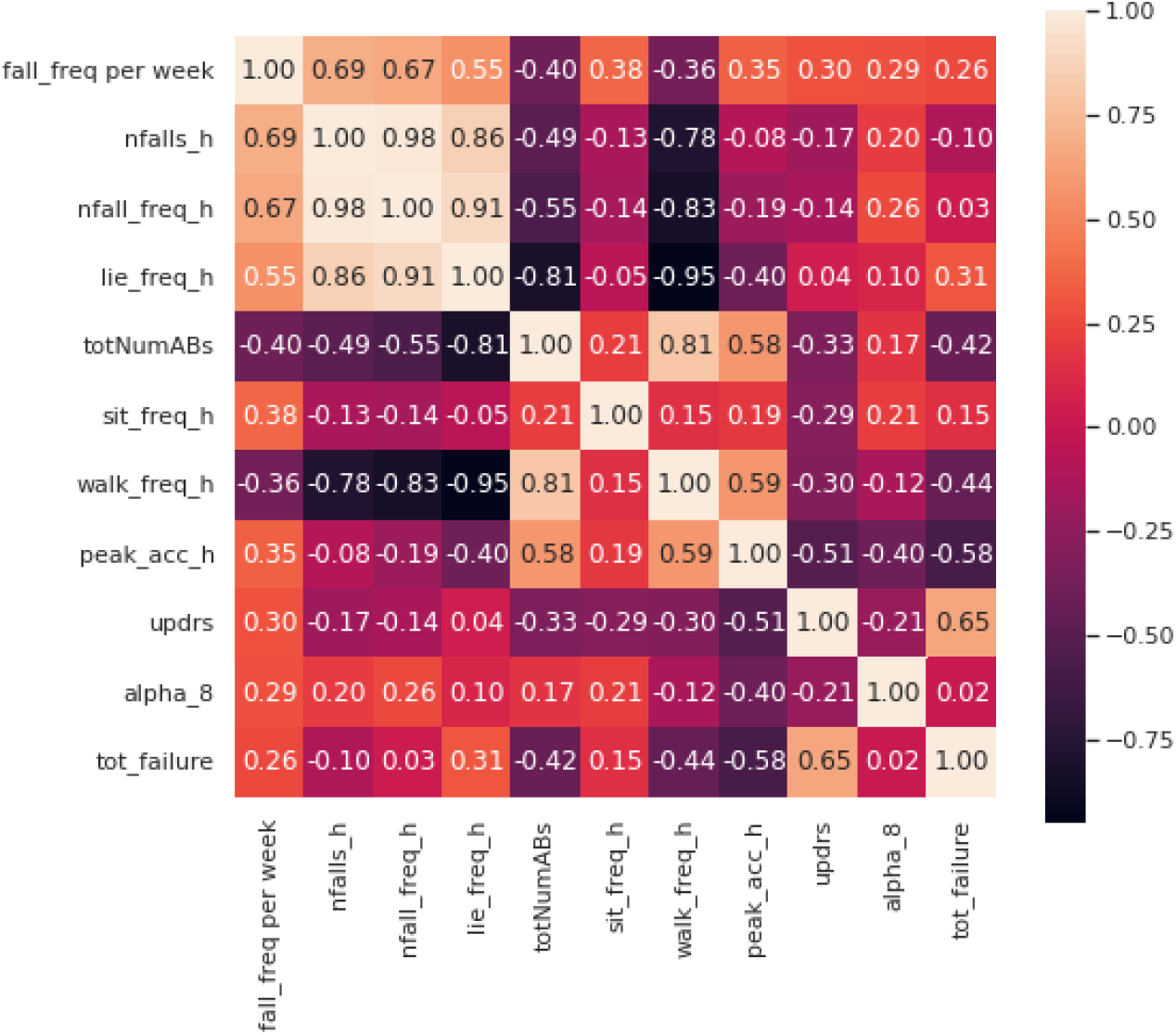
Correlation matrix including the first 10 features with the most correlation with the fall frequency of patients at home.

## 4 Discussion

Using a combination of domain specific knowledge and machine learning techniques, we developed an automatic algorithm for detection and characterization of near-falls and high fall risk activities of the patients. We created a validated, video annotated and quantitative dataset of movement disorder subjects wearing inertial sensors at their home environment. The statistical analysis of our algorithm shows >95% sensitivity in detection of activities apart from near-falls, which showed 80% sensitivity. The correlation analysis of the computed features in our dataset showed that our novel metrics based on near-falls are superior in terms of the highest correlation with patient fall frequency over an entire year of follow-up while clinic-based features were either not correlated or were among the most weakly correlated features.

### 4.1 Video Validation of CNN-LSTM Algorithm in the Home Setting

Using video to validate algorithm predictions based on IMU data is a necessary component to reliably interpret any wearable dataset, but is frequently lacking in studies employing wearables [18]. Studies that do employ some measure of validation typically do so in the clinic or laboratory setting, rather than “in the wild” as we have done in this study [18]. As such, this represents an ecologically valid quantitative dataset that can then be leveraged to understand the factors relevant in producing falls with far more detail compared to the current standard solution which involves qualitative clinical examination or laboratory-based task assessments that may or not be related to real world balance perturbations. Since most falls occur at home and surrounding environment, a wearable dataset that is “validated” in the clinic or lab may not be ecologically valid for fall prediction or other uses. On the other hand, some activities such as walking, standing and bending may be able to be easily validated in the clinic setting and so datasets validated with video “in the wild” should be compared to datasets using clinic-based assessments in order to better understand what can accurately assessed in the clinic vs. which assessments need to occur at home.

Our activity recognition algorithm uses a nonlinear switched-gain observer based on measurements from IMUs worn on leg segments and the chest in order to estimate body segment orientation. The observer estimates the tilt angles and measurement bias is estimated and removed. This has been measured in prior studies using infrared-based motion capture systems to ensure its accuracy [16]. These estimates are then used to train the LSTM deep learning algorithm on all of the activities. Since many of the activity definitions are based on the tilt angles (Supplemental Table 1), this may be one reason that our LSTM activity recognition method showed superior accuracy. In addition, our deep learning network demonstrated lower computation cost compared to the other methods as it reduces the number of raw IMU signals necessary for activity recognition. Future studies should investigate how to reduce both the number of worn IMUs and number of recorded events without affecting diagnostic accuracy in order to minimize the burden on patients wearing the IMUs.

### 4.2 Development of novel behavioral biomarkers of falls

We have developed prospective, predictive falls risk metrics that integrate the patient’s postural response along with data that reflects the patient’s home environment based on near-falls detection. We have included all known gait parameters used in prior studies and current clinical standards in the study, however our proposed metrics showed superior performance in predicting falls in these patients. While there are simple measures which may be more easily measured with a smartphone or smartwatch (e.g., frequency of lying down, number of ambulatory bouts), our study suggests these are inferior to the number or frequency of near-falls. Similarly, clinical tests are typically inadequate in describing the likelihood of the subject falling and in characterizing the extent of their postural instability [7]. Inter- and intra-rater variability in the execution and interpretation of clinical testing likely is responsible for some of their poor predictive power. Furthermore, incidents that trigger stumbles and falls at home are almost certainly different from testing conducted by clinicians or researchers in an artificial environment. Unfortunately, there are few studies on postural instability in home environments and these studies typically do not provide enough validation for their results in real-life situations at home or at least the demonstration of generalizability to the home environment [19]. While there may be overlap in the postural response to balance testing in the clinic or lab and that at home, datasets such as the one described in our study should be used to investigate similarities and differences between these two settings in the future. This could lead to better fall prediction algorithms and improved diagnostic monitoring and treatment evaluations in the clinic, lab and at home in the future. Despite being the most relevant feature of the dataset for fall prediction, near-falls were the most difficult activity to accurately detect with a sensitivity of 80%. This was for several reasons. First, near-falls look similar to other activities (like sit-stand transitions and bending) when examining inertial sensor data. Second, some of the near-falls were so subtle that they could not even be detected in videos. Finally, the natural occurrence of near-falls is relatively rare and obtaining video-validated samples is difficult since most of the patients who are at risk of falling are usually less active or use a walking aid to avoid falling. As such, they were also among the least common events and our algorithm, like all machine learning algorithms, performs better with more samples. Continued data collection with more validated events will likely help increase the accuracy of the algorithm over time.

### 4.3 Future Dataset Usage

We have collected our dataset using an inexpensive wearable system based on inertial sensors to provide kinematic data of PD and NPH patients at home. Typical uses of IMUs worn by movement disorder patients at home are detailed gait analysis and metrics on mobility/ambulation which can be used for a wide variety of purposes such as disease stage assessment, fall prediction, and treatment evaluation, among others [20-23]. An advantage of our system is that it contains data from sensors on the chest and both feet that can be used to give detailed information on the postural response to near falls that occur in a natural setting in addition to all of those typical uses described above. Given the contribution of postural instability to falls in these patients, characterizing postural instability at home could potentially be very useful in their monitoring and treatment evaluation, particularly as their disease progresses and their likelihood of falling increases. Most studies of postural instability are still based on questionnaires or short-term simulations of near-falls in a clinical or lab setup [22]. In addition, while wearable studies are becoming more common, clinical fall risk assessment is usually performed using diaries and questionnaires or one-time evaluations of gait and balance factors of the patients in a clinical trial [2], [3]. These methods are questionable in their quality and credibility due to their short-term and subjective assessment of the patients’ response [7]. Thus, there is a crucial need for a long-term, easily obtained, and objective characterization of gait and postural instability in the home setting as a complement to clinical assessments. We would argue that a dataset such as the one described in this manuscript would represent the first step towards that goal.

One limitation of this dataset is its practicality as the current setup with 5 sensors might not be practical for everyday patient use. Future research should develop algorithms to use as few sensors as possible in optimal locations on the body. Because we were interested mainly in postural instability and falls, we did not include IMUs on the upper limbs and so our dataset does not include hand or arm movements. Given the frequent presence of upper extremity tremor in PD, this is particularly relevant for these patients in their diagnosis, monitoring and treatment evaluation. In addition, many activities of daily living can likely be classified with an upper extremity IMU. Further research should plan to integrate IMU/smartwatch-based data to obtain the widest variety of activities with the best diagnostic accuracy. Wearable usage should also be tailored to the specific usage desired by the clinician and patient. Finally, we plan to further develop activity recognition algorithms using unsupervised and semi-supervised learning methods to increase their accuracy or discover new activities which might have been missed by the current methods.

Even though near-fall detection is difficult to recognize and our algorithm shows 80% sensitivity, near-fall frequency at home was still the most predictive criterion in the linear regression model compared to any other metric. Our results showed that the detection of near-falls is a far more powerful way to examine home monitoring data compared to current methods and should be incorporated into fall prediction algorithms. This validated dataset of movement disorder patients engaged in daily living activities in their homes can serve as a valuable resource for researchers to provide a ground truth for IMU algorithm comparison that include the natural responses of patients at home.

## Data Availability

All data produced in the present study are available upon reasonable request to the authors

## 8 Author Contributions

RAM conceived the original idea. AN developed the algorithms, designed the sensors and measurement setup with the help and supervision of RR. AN, AJ, JH, LS, JJ, TL, ER, YM, SR, KN, CS helped with the data collection and annotations. AN analysed and interpret the results with the help and supervision of RAM. AN and RAM wrote the manuscript with the help of AJ and LS. RAM and RR supervised the project.

## References

[1] T. A. Stoffregen and L. J. Smart Jr, “Postural instability precedes motion sickness,” Brain Res. Bull., vol. 47, no. 5, pp. 437–448, 1998.

[2] C. Colosimo et al., “Task force report on scales to assess dyskinesia in Parkinson’s disease: Critique and recommendations,” Mov. Disord., vol. 25, no. 9, pp. 1131–1142, Jul. 2010, doi: 10.1002/MDS.23072.

[3] J. M. Kane et al., “Revisiting the Abnormal Involuntary Movement Scale: Proceedings From the Tardive Dyskinesia Assessment Workshop,” J. Clin. Psychiatry, vol. 79, no. 3, p. 18344, May 2018, doi: 10.4088/JCP.17CS11959.

[4] C. G. Goetz et al., “Movement Disorder Society-Sponsored Revision of the Unified Parkinson’s Disease Rating Scale (MDS-UPDRS): Scale presentation and clinimetric testing results,” Mov. Disord., vol. 23, no. 15, pp. 2129–2170, Nov. 2008, doi: 10.1002/MDS.22340.

[5] R. A. Hauser, H. Russ, D. A. Haeger, M. Bruguiere-Fontenille, T. Müller, and G. K. Wenning, “Patient evaluation of a home diary to assess duration and severity of dyskinesia in parkinson disease,” Clin. Neuropharmacol., vol. 29, no. 6, pp. 322–330, Nov. 2006, doi: 10.1097/01.WNF.0000229546.81245.7F.

[6] S. Papapetropoulos, “Patient Diaries As a Clinical Endpoint in Parkinson’s Disease Clinical Trials,” CNS Neurosci. Ther., vol. 18, no. 5, pp. 380–387, May 2012, doi: 10.1111/J.1755-5949.2011.00253.X.

[7] A. L. Silva de Lima et al., “Home-based monitoring of falls using wearable sensors in Parkinson’s disease,” Mov. Disord., vol. 35, no. 1, pp. 109–115, Jan. 2020, doi: 10.1002/mds.27830.

[8] F. Motolese et al., “Parkinson’s Disease Remote Patient Monitoring During the COVID-19 Lockdown,” Front. Neurol., vol. 11, p. 1190, Oct. 2020, doi: 10.3389/FNEUR.2020.567413/BIBTEX.

[9] C. Rougier, J. Meunier, A. St-Arnaud, and J. Rousseau, “Robust video surveillance for fall detection based on human shape deformation,” IEEE Trans. Circuits Syst. Video Technol., vol. 21, no. 5, pp. 611–622, May 2011, doi: 10.1109/TCSVT.2011.2129370.

[10] “SparkFun OpenLog Artemis - DEV-16832 - SparkFun Electronics.” https://www.sparkfun.com/products/16832 (accessed Dec. 19, 2021).

[11] “ICM-20948 Datasheet | TDK.” https://invensense.tdk.com/download-pdf/icm-20948-datasheet/ (accessed Dec. 19, 2021).

[12] “Runcam 5 Datasheet,” 2020. https://www.runcam.com/download/runcam5/RunCam5-Manual-EN.pdf (accessed Dec. 19, 2021).

[13] O. Calin, Deep learning architectures. Springer, 2020.

[14] “Extremely Accurate I2C-Integrated DS3231 RTC Datasheet,” 2015 https://datasheets.maximintegrated.com/en/ds/DS3231.pdf (accessed Sep. 06, 2022).

[15] Nouriani, A., McGovern, R., and Rajamani, R., “Nonlinear LMI-based Observer Estimates Improve Accuracy of LSTM Human Activity Recognition Algorithms” Proceedings of Modeling, Estimation and Control Conference (MECC), 2-5 Oct 2022, New Jersey, NJ.

[16] A. Nouriani, R. McGovern, and R. Rajamani. “Step length estimation with wearable sensors using a switched-gain nonlinear observer.” Biomedical Signal Processing and Control 69 (2021): 102822.

[17] Del Din, Silvia, et al. “Analysis of free-living gait in older adults with and without Parkinson’s disease and with and without a history of falls: identifying generic and disease-specific characteristics.” The Journals of Gerontology: Series A 74.4 (2019): 500–506.

[18] Nouredanesh, Mina, et al. “Fall risk assessment in the wild: A critical examination of wearable sensor use in free-living conditions.” Gait & Posture 85 (2021): 178–190.

[19] J. K. Lee, S. N. Robinovitch, and E. J. Park, “Inertial Sensing-Based Pre-Impact Detection of Falls Involving Near-Fall Scenarios,” IEEE Transactions on Neural Systems and Rehabilitation Engineering, vol. 23, no. 2, pp. 258–266, 2015.

[20] I. Pang, Y. Okubo, D. Sturnieks, S. R. Lord, and M. A. Brodie, “Detection of Near Falls Using Wearable Devices: A Systematic Review,” Journal of Geriatric Physical Therapy, vol. 42, no. Lippincott Williams and Wilkins, pp. 48–56, 01-Jan-2019.

[21] N. Mohammadian Rad, T. Van Laarhoven, C. Furlanello, and E. Marchiori, Novelty detection using deep normative modeling for imu-based abnormal movement monitoring in parkinson’s disease and autism spectrum disorders. Sensors, 18(10), p.3533, 2018.

[22] A. Zhan, S. Mohan, C. Tarolli, R. B. Schneider, J. L. Adams, S. Sharma, M. J. Elson, K. L. Spear, A. M. Glidden, M. A. Little, A. Terzis, E. Ray Dorsey, and S. Saria, “Using smartphones and machine learning to quantify Parkinson disease severity the mobile Parkinson disease score,” JAMA Neurology, vol. 75, no. 7, pp. 876–880, Jul. 2018.

