## Supplemental data for "Real world validation of activity recognition algorithm and development of novel behavioral biomarkers of falls in aged control and movement disorder patients"

#### Supplemental Table 1

| Num | Feature | Definition |
|---|---|---|
| 1 | Stand | Standing (chest, thigh and shank angles < 20 deg) |
| 2 | Walk | At least 3 consecutive steps |
| 3 | Sit | Sitting (thigh angles > 45 deg while chest angle < 20 deg) |
| 4 | Stand to sit | Transition from standing/walking to sitting |
| 5 | Sit to stand | Transition from sitting to standing/walking |
| 6 | Turn | Turning in chest at least 45 deg/s |
| 7 | Lie down | Chest tilt angle > 45 deg and Thigh tilt angles > 45 deg |
| 8 | Bend | Bend at least 45 deg while Thigh Angles < 20 deg |
| 9 | Near-Fall | High forward/backward/lateral acceleration (>10 m/s^2) followed by consecutive balancing steps (>1 step) |
| 10 | Fall | Free fall, i.e. when the only force acting on body is gravity and the chest acceleration is about g=9.8 m/s^2 |

#### Supplemental Table 2

| Num | Feature | Definition |
|---|---|---|
| 1 | age | Age of the patient [years] |
| 2 | height | Height of the patient [centimeters] |
| 3 | weight | Weight of the patient [kilograms] |
| 4 | peak_acc | Chest peak acceleration measured in clinical pull tests [meter/second^2] |
| 5 | mean_acc | Chest average acceleration measured in clinical pull tests [meter/second^2] |
| 6 | updrs | MDS-Unified Parkinson's Disease Rating Scale measured in clinic |
| 7 | tot_failure | Total number of failures in clinical pull tests (had to be caught by examiner) |
| 8 | stp_len | Average step length of the patient in clinical pull tests [meters] |
| 9 | rxn_time | Average reaction time of the patient in clinical pull tests [seconds] |
| 10 | rxn_pkAcc_slope | Slope of reaction time vs. peak chest acceleration in clinical pull tests [second^3/meter] |
| 11 | rxn_mAcc_slope | Slope of reaction time vs. mean chest acceleration in clinical pull tests [second^3/meter] |
| 12 | stpl_pkAcc_slope | Slope of step length vs. peak chest acceleration in clinical pull tests [second^2] |
| 13 | stpl_mAcc_slope | Slope of step length vs. mean chest acceleration in clinical pull tests [second^2] |
| 14 | peak_acc_h | Peak chest acceleration measured at home [meter/second^2] |
| 15 | mean_acc_h | Average chest acceleration measured at home [meter/second^2] |
| 16 | stp_len_h | Average step length of the patient measured at home [meters] |
| 17 | Rxn_time_h | Average reaction time of the patient measured at home [seconds] |

| | | |
|---|---|---|
| 18 | rxt_pkAcc_slope_h | Slope of reaction time vs. peak chest acceleration measured at home [second^3/ meter] |
| 19 | rxn_mAcc_slope_h | Slope of reaction time vs. mean chest acceleration measured at home [second^3/ meter] |
| 20 | stpl_pkAcc_slope_h | Slope of step length vs. peak chest acceleration measured at home [second^2] |
| 21 | stpl_mAcc_slope_h | Slope of step length vs. mean chest acceleration measured at home [second^2] |
| 22 | walk_freq_h | Walking frequency at home (total walking duration / total measurement duration) |
| 23 | turn_freq_h | Turning frequency at home (total turning duration / total measurement duration) |
| 24 | bend_freq_h | Bending frequency at home (total bending duration / total measurement duration) |
| 25 | sit_freq_h | Sitting frequency at home (total sitting duration / total measurement duration) |
| 26 | lie_freq_h | Lie down frequency at home (total lying down duration / total measurement duration) |
| 27 | nfall_freq_h | Near-fall frequency at home (total near-falls duration / total measurement duration) |
| 28 | nfalls_h | Total number of near-falls at home |
| 29 | totWalk_Min_day | Total duration of walking at home in each day [minutes/day] |
| 30 | percWalk_day | Percentage of walking at home in each day [%/day] |
| 31 | totNumABs | Total number of ambulatory bouts *see [18] for details |
| 32 | meanABdur | Average ambulatory bouts duration [seconds] |
| 33 | variability | Variability of ambulatory bouts duration [seconds^2] *see [17] for details |
| 34 | alpha | Alpha for ambulatory bouts *see [17] for details |
| 35 | totWalk_Min_day_3 | Total duration of walking (counted only walks of more than 3 seconds) at home in each day [minutes/day] *see [18] for details |
| 36 | percWalk_day_3 | Percentage of walking (counted only walks of more than 3 seconds) at home in each day [%/day] |
| 37 | totNumABs_3 | Total number of ambulatory bouts (counted only ABs of more than 3 seconds) *see [18] for details |
| 38 | meanABdur_3 | Average ambulatory bouts duration (counted only ABs of more than 3 seconds) [seconds] *see [18] for details |
| 39 | variability_3 | Variability of ambulatory bouts duration (counted only ABs of more than 3 seconds) [seconds^2] *see [17] for details |
| 40 | alpha_3 | Alpha for ambulatory bouts (counted only ABs of more than 3 seconds) *see [17] for details |
| 41 | totWalk_Min_day_8 | Total duration of walking (counted only walks of more than 8 seconds) at home in each day [minutes/day] *see [18] for details |
| 42 | percWalk_day_8 | Percentage of walking (counted only walks of more than 8 seconds) at home in each day [%/day] |
| 43 | totNumABs_8 | Total number of ambulatory bouts (counted only ABs of more than 8 seconds) *see [18] for details |
| 44 | meanABdur_8 | Average ambulatory bouts duration (counted only ABs of more than 8 seconds) [seconds] *see [18] for details |
| 45 | variability_8 | Variability of ambulatory bouts duration (counted only ABs of more than 8 seconds) [seconds^2] *see [17] for details |
| 46 | alpha_8 | Alpha for ambulatory bouts (counted only ABs of more than 8 seconds) *see [17] for details |
| 47 | fall_freq_per_week | Fall frequency of the patients at home reported in patient diaries in a week [# falls / week] |

### Supplemental Figures

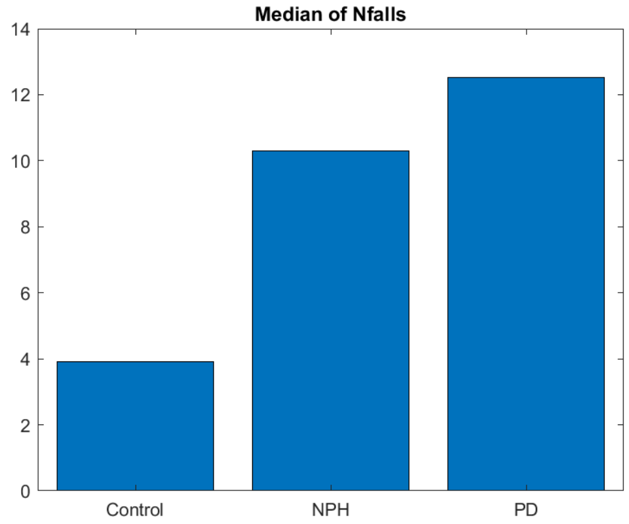


Supplemental Fig 1. Comparison of median of number of near-falls per week for PD vs NPH vs Control subjects.

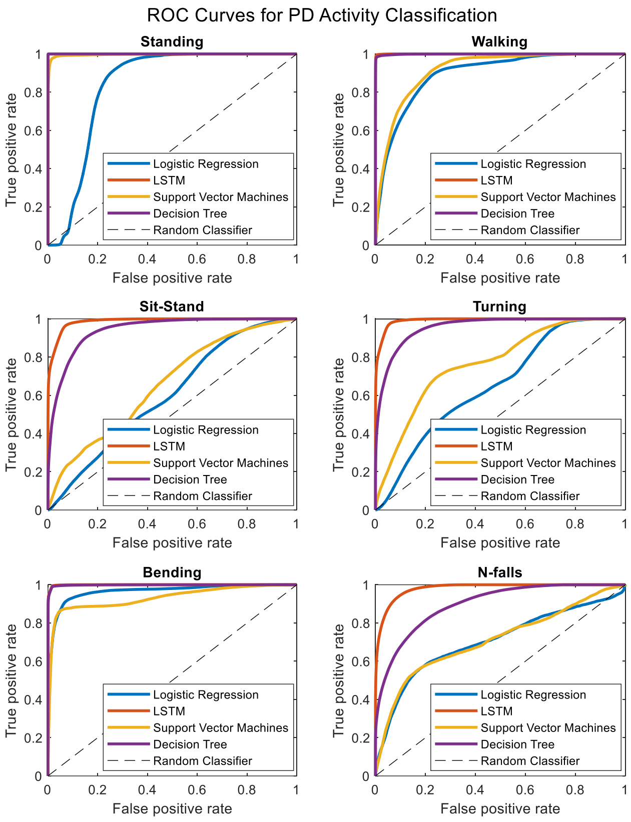


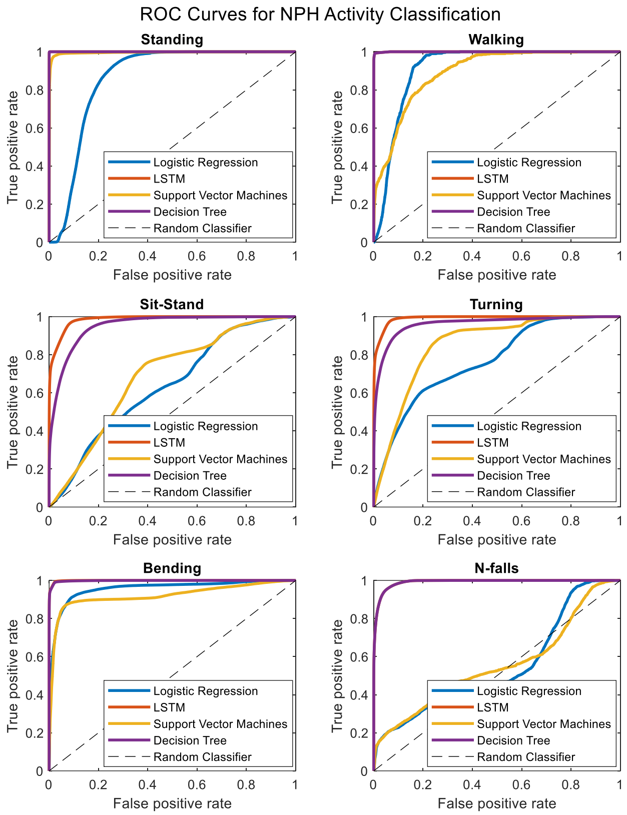


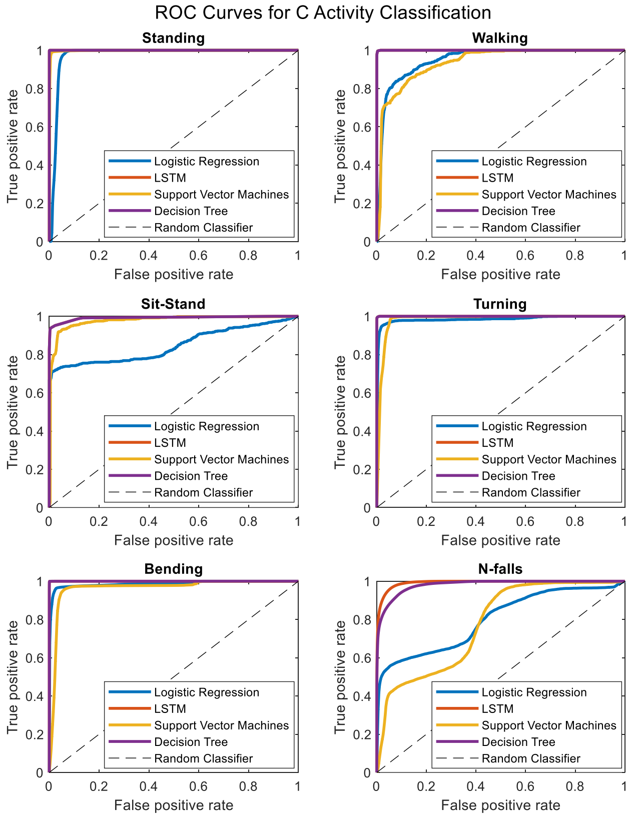


Supplemental Fig 2. Comparison of ROC curves for PD vs NPH vs Control subjects.
